# Second Primary Malignancies in Chronic Lymphocytic Leukaemia; Skin, Solid Organ, Haematological, and Richter’s Syndrome

**DOI:** 10.1101/2021.11.22.21266332

**Authors:** Yandong Shen, Luke Coyle, Ian Kerridge, William Stevenson, Christopher Arthur, Naomi McKinlay, Keith Fay, Christopher Ward, Matthew Greenwood, O. Giles Best, Ann Solterbeck, Alexander Guminski, Stephen Shumack, Stephen P. Mulligan

## Abstract

Chronic lymphocytic leukaemia (CLL) is invariably accompanied by some degree of immune failure. CLL patients have a high rate of second primary malignancy (SPM) compared to the general population. We comprehensively documented the incidence of all forms of SPM including skin cancer (SC), solid organ malignancy (SOM), second haematological malignancy (SHM), and separately Richter’s Syndrome (RS) across all therapy eras. Among the 517 CLL/SLL patients, the overall incidence of SPMs with competing risks were SC 31.07%, SOM 25.99%, SHM 5.19% and RS 7.55%. Melanoma accounted for 30.3% of SC. Squamous cell carcinoma (SCC), including 8 metastatic SCCs, was 1.8 times more than basal cell carcinoma (BCC), a reversal of the typical BCC:SCC ratio. The most common SOM were prostate (6.4%) and breast (4.5%). SHM included 7 acute myeloid leukaemia and 5 myelodysplasia of which 8 were therapy-related. SPMs are a major health burden with 44.9% of CLL patients with at least one, and apart from SC, associated with significantly reduced overall survival. Dramatic improvements in CLL treatment and survival have occurred with immunochemotherapy and targeted therapies but mitigating SPM burden will be important to sustain further progress.

## Introduction

Immune failure associated with chronic lymphocytic leukaemia (CLL) and small lymphocytic lymphoma (SLL) is one of the major and ongoing challenges of the disease. CLL immune failure results in an increased infection risk (1-4), but also a significant rise in the incidence of second primary malignancies (SPM). The advent of immunochemotherapy (ICT) and targeted therapies have significantly improved CLL-related survival, but this enables the emergence of SPMs including skin cancer (SC) and solid organ malignancies (SOM), second haematological malignancies (SHM) such as myelodysplasia (MDS) and acute myeloid leukaemia (AML) and high-grade lymphoid malignancies, i.e., Richter’s Syndrome (RS).

The higher risk of SPM associated with CLL has been recognized for many years, well prior to the modern therapy era (5-10). Manosow and Weinerman (5) in 1975 demonstrated a 3- fold increase of SPM and an 8-fold increase of SC in CLL. Data from M.D. Anderson Cancer Centre (MDACC) in 2009, after the introduction of ICT but prior to the targeted therapy era, showed a 2.2-fold higher risk of SPMs in CLL/SLL compared to the general population (7). In more recent reports, Ishdorj *et al*. (8) in 2019, demonstrated a 4-fold increase in SC with decade-long follow-up of the Manitoba CLL population, while Bond *et al*. (11) in 2020, showed a 2.2-fold higher rate of SPM among CLL patients treated with BTK inhibitors.

As noted by others, SPM risk in CLL has not been systematically addressed (12). In this detailed and comprehensive analysis from a single institution over 40 years, we evaluated all 4 major forms of SPMs assessed as separate entities viz; SC, SOM, SHM, and RS, that occur in CLL (including CLL, SLL, and separately monoclonal B-lymphocytosis [MBL]). Furthermore, we compared the incidence rates of SPM with the CLL literature and with Australian Cancer registry and Australian Institute of Health and Welfare (AIHW) which provides Australian population data for malignancy.

## Methods

### Patient cohort

Data included in this study were obtained from patients managed at Royal North Shore Hospital (RNSH), Sydney, NSW, Australia, with informed consent. Research was approved by the Northern Sydney Local Health District Human Research Ethics Committee (approval number: LNR/14/HAWKE/181). RNSH is the principal hospital for Northern Sydney with a population of ∼1.3 million, the primary source of patients, in addition to which there are specialist referrals to this teaching hospital from across the state of New South Wales (population ∼8 million). All CLL, SLL and MBL patients in the Department of Haematology Audit 4 (S4S) data base were reviewed in detail including every histopathology report and all communications for every patient for the period January 1981 to December 2020. Patients were included in the analysis if there was over one year of follow-up. Patient age was at last follow-up at the time of analysis.

CLL, SLL and MBL patients were diagnosed in accordance with international workshop on CLL (iwCLL) guidelines (13). CLL and SLL were analysed as one disease, hereafter referred to as “CLL”. Follow-up time was calculated from diagnosis of CLL to the last follow-up date. MBL was analysed separately.

All SPMs, including those diagnosed prior to CLL, were summarised in the raw incidence. SCs, including metastatic SC was assessed separately to SOMs. For SHMs, the myeloid malignancies AML or MDS, prior CLL therapy exposed and not, were assessed separately to myeloproliferative neoplasms (MPN). Likewise, low-grade lymphoid malignancies defined as hairy cell leukaemia (HCL), follicular lymphoma (FL), mantle cell lymphoma (MCL), monoclonal gammopathy of undetermined significance (MGUS) and myeloma (MM), were assessed separately. RS followed the standard definition of a “high-grade lymphoid malignancy” in a patient with pre-existing CLL. Mostly in the literature, and our cohort, these were diffuse large B-cell lymphoma (DLBCL), with smaller numbers with Hodgkin’s Lymphoma (HL). Rarely, T-cell variants have been described and we included in the RS group T-cell malignancies of any histology (14, 15).

### Statistical analysis

Statistical analysis and cumulative incidence (and 95% confidence limits) of SPM was determined using the method of Fine and Gray with death as a competing risk. All statistical analyses were performed using SAS software (V14.3). Overall survival was determined using the Kaplan–Meier method and comparisons between specific SPMs and the cohort with CLL only performed using the log-rank test. Hazard Ratios (HR) were calculated using a Cox Proportional hazards model with age group and gender as covariates.

To obtain annual average age-standardised rates (ASR) the crude rate for each age group in 5-year age bracket was determined as the number of individuals with malignancy of interest/total person years, multiplied by 100,000 (to express rate as number per 100,000). The crude rate was then multiplied by the proportion of individuals in that age group according to Australian Bureau of Statistics (ABS) data (2001 Australian standard population). The overall average annual age adjusted rate (per 100,000) was the sum of the adjusted rates for each age group. This was compared with data provided by the Australian Institute of Health and Welfare (AIHW), an independent statutory Australian Government agency that provides health-related information and statistics (aihw.gov.au) (16). Standardised incidence ratio (SIR) was calculated as the ASR for our cohort/ASR from the AIHW dataset. Confidence limits (CL) were calculated using the methods described by Rothman and Greenland (17).

## Results

### Patient characteristics

A total of 570 patients diagnosed with CLL (501), SLL (16) or MBL (53) from January 1981 to December 2020 with a minimum 1-year follow-up were included. Median follow-up duration was 11 years. The male to female (M:F; 323:194) ratio of the CLL cohort was 1.7:1 with a median age of 74.0, and age range of 18 to 101 years. The follow-up time of CLL ranged from 1 to 39 years, during which 122 patients recorded a SC (23.6%) and 103 recorded a SOM (19.9%), respectively. There were 30 cases with a SHM, not including 31 RS discussed separately. Overall, 232 of 517 (44.9%) CLL patients had at least one SPM (Table 1) (including SPMs diagnosed prior to the CLL follow-up period).

**Table 1.** Clinical features of CLL and MBL patients. ^*^ indicates patients with at least one or more diagnosis of either skin, solid organ malignancies, second haematological malignancies or Richter’s syndrome, in addition to their CLL or MBL. Patients diagnosed with multiple types of SPMs were counted as separate incidences. Median age was calculated at the last follow-up or the age when patient died.

|  | Number of patients | Male : Female | Median age at the last follow-up (inter-quartile) | Mortality |
| --- | --- | --- | --- | --- |
| **<u>CLL cohort</u>** | 517 | 1.7 : 1 | 74.0 (66.0-83.0) | 15.5% |
| CLL only | 285 (55.1%) | 1.5 : 1 | 73.0 (62.0-82.0) | 11.9% |
| Skin cancer | 122 (23.6%) | 2.0 : 1 | 77.5 (70.3-84.0) | 14.8% |
| Solid organ malignancies | 103 (19.9%) | 2.0 : 1 | 76.0 (69.0-84.0) | 19.4% |
| Haematological malignancies | 30 (5.8%) | 2.3 : 1 | 75.0 (66.0-80.8) | 43.3% |
| Richter's Syndrome | 31 (6.0%) | 2.4 : 1 | 68.0 (63.3-73.0) | 35.5% |
| Any SPMs * | 232 (44.9%) | 1.9 : 1 | 76.0 (68.8-84.0) | 19.8% |
| **<u>MBL cohort</u>** | 53 | 1.6 : 1 | 75.5 (67.0-82.0) | 5.7% |
| MBL only | 36 (67.9%) | 1.8 : 1 | 75.5 (67.0-80.5) | 5.6% |
| Skin cancer | 9 (17.0%) | 1.3 : 1 | 84.0 (75.0-92.0) | 11.1% |
| Solid organ malignancies | 75 (13.2%) | 0.4 : 1 | 78.0 (73.0-81.5) | 0.0% |
| Any SPMs * | 17 (32.1%) | 1.8 : 1 | 78.0 (67.3-84.8) | 5.9% |

Of the 517 CLL patients, *IGHV* status was available on 38 (7.4%, 27 mutated and 11 unmutated) and cytogenetics analysis on 106 patients (20.5%). Of the 11 unmutated patients, there were 5 SCs, 4 SOMs, 1 myeloma and no RS recorded; of the 27 mutated patients, there were 6 SC, 4 SOMs and no SHM or RS.

A total of 216 patients had at least one line of treatment. Of first-line treatments, 66 were alkylating/chlorambucil based, 125 fludarabine, cyclophosphamide, rituximab (FCR, or other fludarabine) based, 10 targeted therapies, and 15 others not specified (Supplementary Figure 1). Most targeted therapy (68 patients) was for relapsed CLL. Of the 216 treated patients, 106 (49.1%) had at least one form of SPM, and 63 of 106 (29.2% of treated patients) developed a SPM 1.5 years (median) after treatment for their CLL; 24 had a SPM before CLL treatment; and in 19 the sequence was unclear.

There were 53 patients with MBL (Table 1) with a median age of 75.5 years, and M:F ratio of 1.6:1 (33:20). Of the 53 MBL patients, 9 (17.0%) had SC (5 melanoma, 7 NMSC, including 2 BCC, 3 SCC, 1 sebaceous cell carcinoma and 1 unclassified); and 7 (13.2%) had SOMs, 2 with prostate cancer, and 1 each of breast, bladder, thyroid (papillary cell), pancreas and colon. There were 2 MBL patients with DLBCL consistent with RS and 2 diagnoses of MBL with MGUS.

### Second malignancy in the CLL cohort

#### Skin cancer

Of the 517 CLL patients, 122 had a SC including those diagnosed prior to CLL (Table 2). Among these 122 patients, 18 had SC prior to the diagnosis of CLL with a median age 69.0 years, 59 were diagnosed after the CLL with median age of 62.0 years, and 30 patients could not be chronologically traced (Supplementary Figure 2). Melanoma accounted for 37 (30.3% SCs; M:F ratio 1.5:1) of the 122 SC patients, while there were 101 (82.8% SCs; M:F ratio 2.4:1) non-melanoma skin cancers (NMSCs) (basal cell carcinoma [BCC] and squamous cell carcinoma [SCC]) (16 had both melanoma and NMSC, with 14 NMSCs not classified). We observed 33 patients with BCC (M:F ratio 0.9:1) and 58 with SCC (M:F ratio 3.5:1). Metastatic SCC occurred in 8 (13.8% SCC; 6.6% SCs). 2 patients had Merkel cell carcinoma (2.0% NMSC; 1.6% SCs) and 3 had CLL skin infiltration; the latter are included to indicate relative incidence but are not included in the “skin cancer” analysis (Table 2).

**Table 2.** Skin cancers and solid organ malignancies in the CLL cohort. Numbers in this table indicate all CLL patients with skin cancers or solid organ malignancies including those diagnosed prior to their CLL. NMSC incidences included BCC, SCC, Merkel cell carcinoma and cases of unclassified NMSC cases. Patients diagnosed with multiple types of skin cancers were counted as separate incidences. NMSC: non-melanomatous skin cancer; BCC: basal cell carcinoma; SCC: squamous cell carcinoma. ^*^Sebaceous cell carcinoma was identified in a MBL patient and is not included in the CLL analysis cohort; ^**^CLL infiltration not included in “skin cancer” statistics and provided only for comparison. ^***^UPC: uncertain primary carcinoma.

| **<u>Skin Cancers</u>** | Cases | % of CLL cohort | Male |  | Female |  |
| --- | --- | --- | --- | --- | --- | --- |
|  |  |  | Cases | % in male (81) | Cases | % in female (41) |
| Melanoma | 37 | 7.2% | 22 | 27.2% | 15 | 36.6% |
| NMSC | 102 | 19.8% | 72 | 88.9% | 30 | 73.2% |
| BCC | 33 | 6.4% | 16 | 19.8% | 17 | 41.5% |
| SCC | 58 | 11.2% | 45 | 55.6% | 13 | 31.7% |
| Merkel | 2 | 0.4% | 1 | 1.2% | 1 | 2.4% |
| Sebaceous cell carcinoma* | 1 | 0.2% | 0 | 0.0% | 1 | 2.4% |
| CLL skin infiltration** | 3 | 0.6% | 3 | 3.7% | 0 | 0.0% |
| **<u>Solid organ malignancies</u>** | Cases | % of CLL cohort | Cases | % in male (70) | Cases | % in female (33) |
| Breast | 23 | 4.5% | 0 | 0.0% | 23 | 69.7% |
| Lung | 12 | 2.3% | 8 | 11.4% | 4 | 12.1% |
| Colorectal | 11 | 2.1% | 8 | 11.4% | 3 | 9.1% |
| Kidney | 5 | 1.0% | 4 | 5.7% | 1 | 3.0% |
| Endocrine (pituitary, thyroid, parathyroid) | 5 | 1.0% | 3 | 4.3% | 2 | 6.1% |
| Bladder | 4 | 0.8% | 4 | 5.7% | 0 | 0.0% |
| Pancreas | 3 | 0.6% | 3 | 4.3% | 0 | 0.0% |
| Oesophagus | 3 | 0.6% | 3 | 4.3% | 0 | 0.0% |
| Schwannoma | 3 | 0.6% | 3 | 4.3% | 0 | 0.0% |
| Brain (glioma, meningioma) | 2 | 0.4% | 1 | 1.4% | 1 | 3.0% |
| Testicular | 2 | 0.4% | 2 | 2.9% | 0 | 0.0% |
| Ovarian | 1 | 0.2% | 0 | 0.0% | 1 | 3.0% |
| Endometrial | 1 | 0.2% | 0 | 0.0% | 1 | 3.0% |
| Chondrosarcoma | 1 | 0.2% | 1 | 1.4% | 0 | 0.0% |
| Lymph node metastasis (UPC***) | 1 | 0.2% | 1 | 1.4% | 0 | 0.0% |

A total of 57 patients with SC were treated for their CLL, 26 (21.3% SCs) before the diagnosis of SC (4 chlorambucil based, 11 fludarabine based, 8 targeted therapies, and 3 others); 12 had no CLL treatment prior to SC diagnosis (9.9% SCs); and 19 could not be chronologically traced. The median time of between treatment of CLL and diagnosis of SC was 18 months.

#### Solid organ malignancies

There were 103 patients diagnosed with one or more SOMs including those diagnosed prior to their CLL (Table 2). Prostate and breast cancers were the most common SOMs in the CLL cohort, 33 prostate cancers (47.1% male SOMs) and 23 breast cancers (69.7% female SOMs), respectively. Lung and colorectal cancers ranked third and fourth. Details of other SOMs and the diagnosis time relative to the CLL are shown in Table 2 and Supplementary Figure 3.

Of the 103 patients, 19 (18.4% SOMs) were treated for their CLL (4 chlorambucil based, 8 fludarabine based, 5 targeted therapies, and 2 others) prior to the diagnosis of SOMs. Median time between those treatments and diagnosis of SOMs was 12 months.

#### Secondary haematological malignancies Myeloid neoplasms

AML and MDS were diagnosed in 7 and 5 patients respectively (Table 3). Of the 7 AML cases, 4 were previously treated, 2 with FCR, and of the 5 MDS cases, 4 were CLL treated, 3 with FCR, totalling 8/12 “therapy-related” (66.7%) AML/MDS (t-AML/MDS). Of the 124 CLL patients treated with FCR (not including other fludarabine-based therapy) in any line of treatment, 5 developed t-AML/MDS (4.0%, [or 3.1% with all fludarabine-based therapy included]). Treatment history of the 12 AML/MDS patients was summarised in Supplementary Table 1.

**Table 3.**
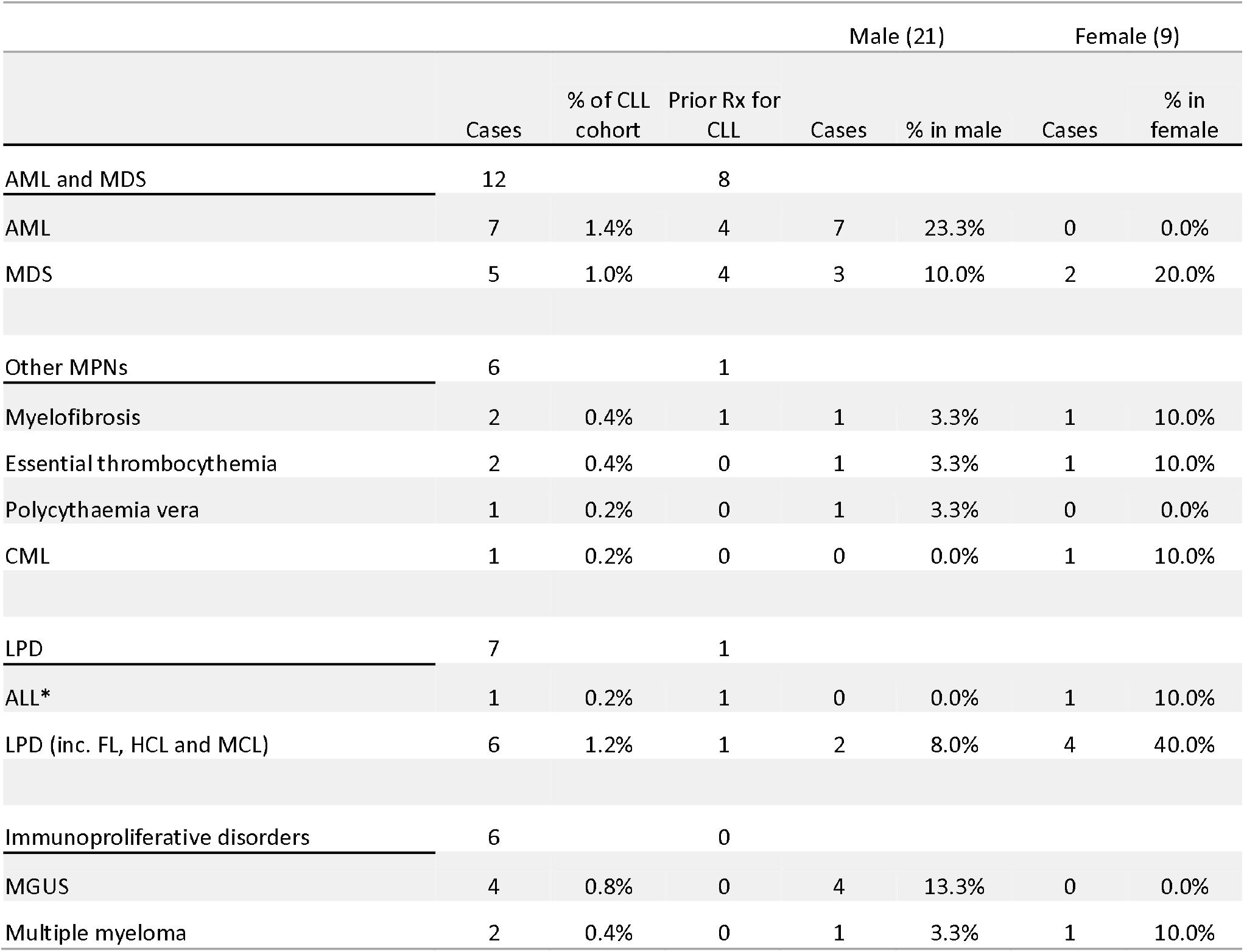
Second haematological malignancies in the CLL cohort. Numbers in this table indicate all CLL patients with second haematological malignancies including those diagnosed prior to their CLL. MPN: myeloproliferative neoplasm; AML: acute myeloblastic leukaemia; MDS: myelodysplastic syndromes; CML: chronic myeloid leukaemia; LPD: low-grade lymphoproliferative disorder; ^*^ALL: acute lymphoblastic leukaemia; FL: follicular lymphoma; HCL: hairy cell leukaemia; MGUS: monoclonal gammopathy of undetermined significance. ^*^1 case of ALL included in the RS analysis, not counted as “LPD” but included for comparison with other SHM. ^*^See Supplementary data regarding the ALL patient.

All 12 cases of AML or MDS were diagnosed after the CLL at a median time of 6 and 10 years, respectively. The “therapy-related” 4 AML and 4 MDS occurred at median time from CLL therapy of 3 and 3 years, respectively. The median survival (calculated from the time of AML or MDS diagnosis) after AML or MDS diagnosis was 4 and 24 months, respectively. To date, 2 patients have developed AML after targeted therapy although follow-up is much shorter (median 36 months), and both had received prior immunochemotherapy. The 3 AML cases with no prior CLL therapy occurred 4, 7, and 8 years after the diagnosis of CLL; survival after AML diagnosis was 1, 1, and 4 months between the age of 60-80 years. The 1 MDS case with no prior CLL therapy (in his 80s) was alive at study cut-off date, 36 months after MDS diagnosis.

MPN was diagnosed in 6 patients (1.2%) including 2 idiopathic myelofibrosis (MF), 2 essential thrombocythemia (ET), 1 polycythaemia vera (PV), and 1 chronic myeloid leukaemia (CML) (Table 3). Of these MPNs, 2 MF, 1 PV and 1 CML were diagnosed after the CLL, and 1 MF patient was previously CLL treated with FCR 1 month earlier. The 2 cases of ET were diagnosed prior to CLL with no prior treatment for the ET.

#### Other lymphoproliferative and plasma cell disorders

There were 6 patients (2 FL, 3 MCL, and 1 HCL) diagnosed with a second lymphoproliferative disorder (Table 3). Both FLs were diagnosed before the CLL, including an unusual concurrently diagnosed combination of 3 haematological malignancies: SLL and FL in a lymph node biopsy (documented distinct by histology and immunohistochemistry), and DLBCL (hence RS) in a gastric biopsy. Diagnosis of CLL and HCL was made concurrently in a female in her 60s, from blood morphology and distinctive immunophenotypes. There were 3 CLL patients with confirmed MCL by phenotype and genetics, all males aged in their 40s, 50s, and 60s at CLL diagnosis. One (in his 40s) was diagnosed concurrently and the other 2 after the CLL. The male (in his 50s) was then diagnosed with MCL, and then AML 5 years later and died 1 month later (hence included in myeloid neoplasm analysis).

There were 6 CLL patients with plasma cell dyscrasias, 4 MGUS and 2 MM. Of these, 4 (3 MGUS and 1 MM) were diagnosed within 6 months after the CLL. The other 2 were diagnosed with MGUS or MM 13 years after CLL. One patient with MGUS and 1 MM were treated with obinutuzumab plus chlorambucil and FCR respectively for their CLL, after diagnosis of MGUS/MM.

#### Richter’s syndrome

RS occurred in 31 patients including 22 DLBCL (73.3% of RS), and 4 Hodgkin’s Lymphoma (HL; 13.3% of RS). We included all high-grade lymphoid malignancies, both of B-cell type (2 B-cell prolymphocytic leukaemia [B-PLL], 1 B-ALL, and 2 T-cell malignancies (1 aggressive peripheral T-cell lymphoma-NOS [not otherwise specified]), and 1 subcutaneous panniculitis-like T-cell lymphoma) (Table 4). The 1 ALL patient was not on lenalidomide (See Supplementary Table 2). RS was diagnosed at a median 6 years after the CLL, and 1.5 years (median) after the last CLL treatment. The median survival (from onset of RS) after RS diagnosis was 24 months.

**Table 4.** Richter’s syndrome. Table includes 2 MBL patients with RS. Numbers in this table indicate all CLL patients with Richter’s syndrome including those diagnosed prior to their CLL. DLBCL: diffuse large B-cell lymphoma; ALL: acute lymphoblastic leukaemia. ^*^Duration from CLL diagnosis (Dx) indicates the median time between diagnosis of CLL and RS.

|  |  |  |  | Male (22) |  | Female (9) |  |
| --- | --- | --- | --- | --- | --- | --- | --- |
|  | Cases | % of CLL cohort | Duration from CLL Dx* | Cases | % in male | Cases | % in female |
| DLBCL | 22 | 73.3% | 6 yrs. | 17 | 81.0% | 5 | 55.6% |
| Hodgkin's lymphoma | 4 | 13.3% | 9 yrs. | 2 | 9.5% | 2 | 22.2% |
| B-cell prolymphocytic leukaemia | 2 | 6.7% | 4.5 yrs. | 2 | 9.5% | 0 | 0.0% |
| T-cell lymphoma | 2 | 6.7% | 6.5 yrs. | 1 | 4.8% | 1 | 11.1% |
| B-ALL* | 1 | 3.3% | 11 yrs. | 0 | 0.0% | 1 | 11.1% |

### Overall Survival and competing risk analysis

The overall survival (OS) of CLL patients with and without any SPM was illustrated by the Kaplan–Meier curves (Figure 1). OS was worse with SOM (HR 1.89, *p=*0.0195), RS (HR 3.42, *p=*0.0003) and AML/MDS (HR 7.44, p<0.001), but not for SC (HR 1.2, *p=*0.5420).

**Figure 1.**
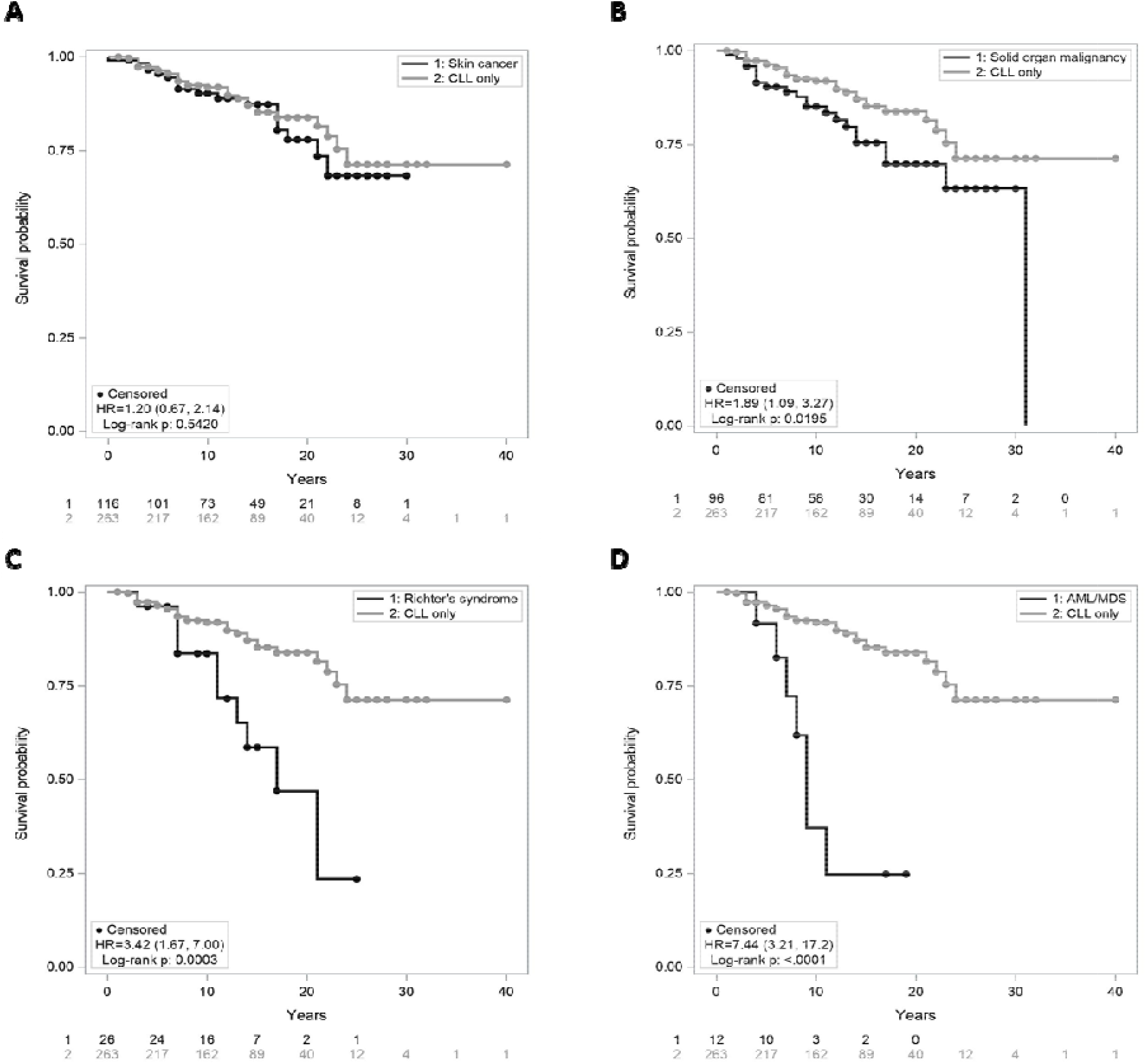
Overall survival (OS) of CLL patients with a second primary malignancy. The survival time was calculated from the time of the CLL diagnosis. (A) analysis by skin cancer; (B) analysis by solid organ malignancies; (C) analysis by Richter’s syndrome; (D) analysis of AML and MDS. P-values were calculated using the Log-Rank tests. P-values < 0.05 were considered statistically significant. Numbers at risk were listed for each panel under the x-axis.

The occurrence of death before SPM can bias analyses of incidence. Therefore, calculation of overall incidence of SPMs in the CLL cohort was adjusted using death as a competing risk (Table 5). This analysis only considered incidences of SPMs developed after the diagnosis of CLL and excluded patients with missing key clinical information. The adjusted overall incidence of all SPM was 59.36%, SC was 31.07%, SOM was 25.99%, RS was 7.55%, SHM was 5.19% and AML/MDS was 2.71%. Cumulative incidence rates of SPMs by age groups at CLL diagnosis was higher in younger age groups for all types of SPMs except for SHM (Supplementary Figure 4), i.e. a higher risk of SPM during their longer time course of CLL.

**Table 5.** Summary of SPM with death as a competing risk. Table summarises the overall incidence of SPMs in the CLL cohort using death as a competing risk. This analysis included SPMs diagnosed after the diagnosis of CLL only, and excluded patients with incomplete information, hence discrepancies between raw incidence numbers and total numbers in the analysis.

| Type of malignancy | Number developing SPM | Number dying before SPM | Number alive and disease free | Total number in analysis | Overall incidence (95% confidence limits) |
| --- | --- | --- | --- | --- | --- |
| Any malignancy | 144 | 31 | 232 | 407 | 59.36% ( 47.36% , 69.49% ) |
| Skin cancer | 70 | 59 | 305 | 434 | 31.07% ( 22.76% , 39.72% ) |
| Solid organ malignancy | 47 | 55 | 329 | 431 | 25.99% ( 13.71% , 40.10% ) |
| Richter's syndrome | 24 | 67 | 386 | 477 | 7.55% ( 4.76% , 11.18% ) |
| Haematological malignancy | 19 | 66 | 385 | 470 | 5.19% ( 3.16% , 7.94% ) |
| AML/MDS | 9 | 70 | 398 | 477 | 2.71% ( 1.31% , 4.98% ) |

Analysis was performed to evaluate potential causal relationship between CLL therapy and SPM development. There was no treatment effect for SC (HR 1.16, CL 0.72, 1.87) or SOM (HR 0.95, CL 0.52, 1.71). RS (HR 2.18, CL 0.95, 5.01) did appear to have a treatment effect with “any prior CLL therapy”, and a marked effect with fludarabine therapy (HR 7.42, CL 0.92, 60.11). There appeared to be a treatment effect for AML/MDS (HR 3.15, CL 0.66, 15.01), but when fludarabine was compared to “any other form of CLL therapy”, this effect was lost (Supplementary Figure 5). Both the RS and AML/MDS numbers were small, and these effects need to be examined on larger numbers of patients.

### Age standard incidence of SPM for CLL compared to the Australian population

The age standard incidence (ASR) for the CLL and MBL cohort was calculated as described in Methods. The RNSH North Sydney Area closely matches the general Australian population (Supplementary Figure 6). The ASR of SPMs for CLL and MBL (Table 6) were all substantially higher than the Australian population. AML and MDS had relatively small numbers resulting in wide confidence intervals.

**Table 6.**
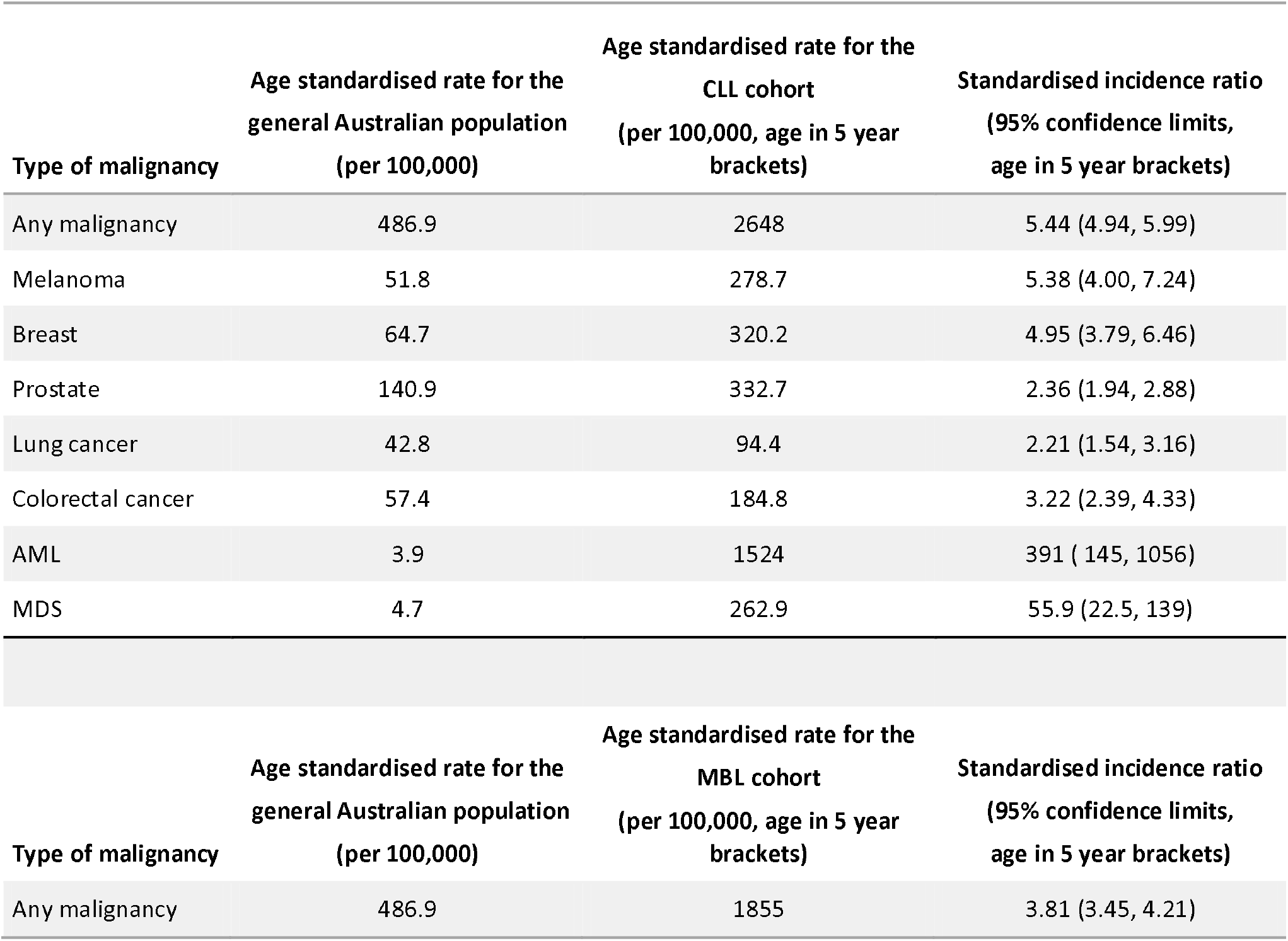
Summary of age adjusted rates using data for 2015 from AIHW report cancer in 2019, age adjustment using 2001 population data and comparison with the CLL and MBL cohort. Age standard rates (ASR) for the CLL cohort was calculated using methods described in the statistical analysis section. ASR for the general Australian population was obtained from the Australian Institute of Health and Welfare (AIHW) caner in Australian 2019 report. ASR was calculated for both 10- and 5-year brackets with minimal differences, age in 5-year brackets was shown.

## Discussion

The development of a SPM has a strong association with CLL. This association long pre-dates the advent of ICT and targeted therapy but appears to be a more significant clinical issue in the current era as patients survive much longer with their CLL. Following the introduction of ibrutinib in Australia in 2012 with the RESONATE trial (18), our centre observed, for the first time, higher mortality due to SC instead of CLL (19).

In the context of skin cancer, melanoma is mandated as a cancer registry reportable malignancy in Australia, while NMSCs (BCCs and SCCs) numbers are only reflected by surveys and insurance records. Skin cancer in a 2009 audit at our institution and an NSW coastal centre was extremely common with NMSC in 58.9% (96/163) with differences between metropolitan versus regional centres (36.7% vs 62.8%), likely reflecting lifestyle, occupation and different UV radiation. The incidence of melanoma was identical (9.8%). In this study, 23.6% of CLL patients had SC, with melanoma in 37 (7.2% of CLLs) and NMSC in 102 (19.8% of CLLs). Hence, the incidence of melanoma is comparable between 2009 and 2020 (9.8% vs 7.2% respectively), while the incidence of NMSC is lower (36.7% vs 19.8% respectively). This period corresponds with long-term results from sun-exposure education campaigns and suggest that NMSC risk in CLL can be modified. Comparison with a cohort of CLL patients from Manitoba, Canada, undergoing regular dermatological evaluation, showed comparable competing risk adjusted incidence of SC (31.07% vs 28.0%) (8).

NMSC is not a reportable malignancy in Australia, and indeed nor in many cancer registries. Staples *et al*. (20) showed the number of SCC in the Australian population was 2-times lower than BCC. By contrast, the incidence rate of SCC in our CLL cohort was 1.8-times higher than BCC. This inversion of the SCC:BCC incidence rates in CLL is highly consistent with solid organ transplant patients on prolonged immunosuppression (21) and emphasizes immune failure contributing to NMSC risk. Furthermore, metastatic SCC (met-SCC) occurred in 8 patients (of 58 SCC; 13.8%). Comparison with the New Zealand (NZ) general community (with similar UV levels) reported 1.9-2.6% met-SCC (22), while a NZ CLL cohort (23) showed 9.9% (6/61) met-SCC. Velez *et al*.(24) at 2 Boston academic centres had 10.7% (3/28) met-SCCs over 20 years, demonstrating highly comparable met-SCC rates in CLL in Australia, NZ and the USA, again supporting a role of immune failure.

Melanoma was the fourth most common cancer in Australia in 2019 with an ASR of 51.8 (25) but substantially higher in our CLL cohort (ASR 278.7, SIR 5.38, CL 4.0, 7.2, Table 6). Australian melanoma incidence rises with age with ASR from 80.2 for age 50-59 years to 229.1 over 80 years (26). Similarly in CLL, both melanoma and NMSC rates rise with age with both substantially higher than general population highlighting both age (20) and CLL-related immune failure as contributing factors for SC in CLL. In younger patients, we observed a higher relative incidence of SPM again consistent with immune failure (Supplementary Figure 4).

SOMs were observed in 103 of the 517 CLL cohort. Consistent with the literature (12), prostate and breast cancer were the two largest contributors of SOMs; 47.1% of male cancers were prostate, and 69.7% of female cancers were breast cancers. AIHW in 2019 showed a community ASR of 140.9 for prostate and 64.7 for breast cancer. In our CLL cohort the ASR was 2.46-times higher (CL 2.02, 2.99) for prostate (346.4) and 4.66-times higher (CL 3.56, 6.10) for breast cancer (320.2) (Table 6). Patients with SOMs had significantly higher mortality than patients with CLL only (*p*=0.0195) with over 1.89-fold higher risk of death (Figure 1), consistent with the literature (27). The MBL cohort also showed comparable incidence rates for prostate and breast cancer. In CLL, lung and colorectal cancers were likewise 2.79-times (CL 1.97, 3.96) higher (ASR 119.6 vs 42.8) and 3.94-times (CL 2.95, 5.26) higher (ASR 225.9 vs 57.4) than the Australian population, respectively (25).

Focusing on AML and MDS, the 2.71% competing risk adjusted incidence (12/517) was higher in our CLL cohort compared to 0.6% by Lenartova *et al*. (28) in the Norwegian CLL registry. Australian community risk of AML to age 75 is 1 in 390 (0.26%) (29). In our CLL cohort, AML total numbers, therapy-related AML, and de novo AML at 7 (1.4%), 4 (0.8%) and 3 (0.6%) respectively are higher than population (Table 6 ASR AML in CLL 1524 vs 3.9 population, SIR 391, CL 145, 1056). Our incidence of t-AML/MDS at 4.0% (5/124), was very similar to the MDACC data^7^ (5.1%; 12/234) calculated in the same manner. In our cohort, 3/7 AML occurred with no prior CLL therapy. The Danish CLL registry of 4286 patients recorded 40 (0.93%) acute leukaemia, and 34 (85%) had no prior therapy (10). They found a fludarabine risk for MDS, but not acute leukaemia (only 2/6 treated received fludarabine) (10). Our data suggest an “any CLL therapy” effect but not a specific fludarabine effect (Supplementary Figure 5), again with the important caveat of very low numbers. Our data and the Danish registry suggest that CLL itself represents a risk for AML (RNSH 0.6% untreated CLL vs 0.26% Australian population; Danish 0.79% untreated CLL vs 0.15% in ‘214,150 comparators’). Given how frequently AML risk is invoked as a reason to avoid FCR, this is an important issue to address in large CLL cohorts with treated and untreated patients. The coexistence of CLL and MPN in our cohort (1.2%) is similar to Italian (∼1%) (30) but higher than Danish data (0.16%).

RS occurred in 31 patients (7.55% adjusted incidence) (including 2 DLBCL within the 53 MBL), consistent with the incidence of RS in recent publications (31-33). Over 70% of cases were DLBCL, with smaller numbers of HL and T-NHL. Over half (17/30) had received treatment for their CLL, and 16 of those 17 were with FCR. Rossi *et al*. (34) and Parikh *et al*. (33) also reported over half of their RS patients were CLL treated. Statistics from MDACC showed the risk of RS following FCR was 6.6% (35), while the German CLL8 trial showed the risk was higher in the FC (6.3%) than the FCR arm (3.2%) (31). Analysis using death as competing risk, the HR of RS in treated CLL versus untreated was 2.18 (CL 0.95, 5.01, Supplementary figure 5). There is insufficient evidence, due to small sample sizes and wide confidence intervals to determine whether RS is associated with fludarabine. Future investigation with larger sample sizes is warranted.

The comprehensive analysis of all forms of SPMs over a very long follow-up is a strength of this study. It accurately reflects the natural history of CLL in a large, mainly community-based, single institution cohort with relatively uniform management. Very long follow-up has potential ascertainment bias by selecting patients with more indolent CLL, and hence longer survival, perhaps enabling more of these patients to develop SPMs. CLL prognostic markers such as chromosomal mutations and IGHV status were available in a small proportion having become available late during this 40-year long timeframe; in any event, staging and karyotype are dynamic factors that change with time. CLL patients when diagnosed with an SOM were usually referred to an oncologist occasionally limiting our ability to precisely identify the cause of death, but most subsequent mortality appeared due to the SOM rather than CLL.

The high incidence of SPMs (ASR 2648 in CLL versus 486.9 in population) has a substantial impact and health burden upon CLL patients, their management and survival. Routine monitoring of the skin and education to avoid sun exposure are essential for CLL patients, as data suggests these measures lower NMSC risk. Education and surveillance for SOM are also vital as these significantly shorten survival, and early detection has at least the potential to improve outcomes. Hence adherence to breast and prostate cancer surveillance guidelines, cessation of smoking, evaluation of iron deficiency for potential gastro-intestinal tract malignancy, and investigation of suspicious symptomatology in other organ systems, are important. As SPMs occur in over half of all CLL patients, the frequent exclusion of such patients in clinical trials limits our understanding of “real-world” outcomes. CLL has seen dramatic improvements in treatment and survival since the introduction of ICT and targeted therapies, and the high incidence and health burden of SPMs may be a key limitation to further progress in this disease.

## Data Availability

All data produced in the present work are contained in the manuscript

## Author contributions

SM, LC, IK, WS, CA, NM, KF, CW, MG and SS collected the data. SM and SS confirmed the accuracy of the SC data. YS compiled the data for analysis and wrote the first draft of the manuscript. AS performed the statistical analysis. YS and SM interpreted the data and prepared the final manuscript, which all authors reviewed and approved.

## Acknowledgements

Dr Kevin Phan, Dermatology RNSH, provided additional SCC references.

## Data Availability Statement

The data that support the findings of this study are available on request from the corresponding author. The data are not publicly available due to privacy or ethical restrictions.

