## Supplemental tables/figure for "Second Primary Malignancies in Chronic Lymphocytic Leukaemia; Skin, Solid Organ, Haematological, and Richter’s Syndrome"

### Slide 1
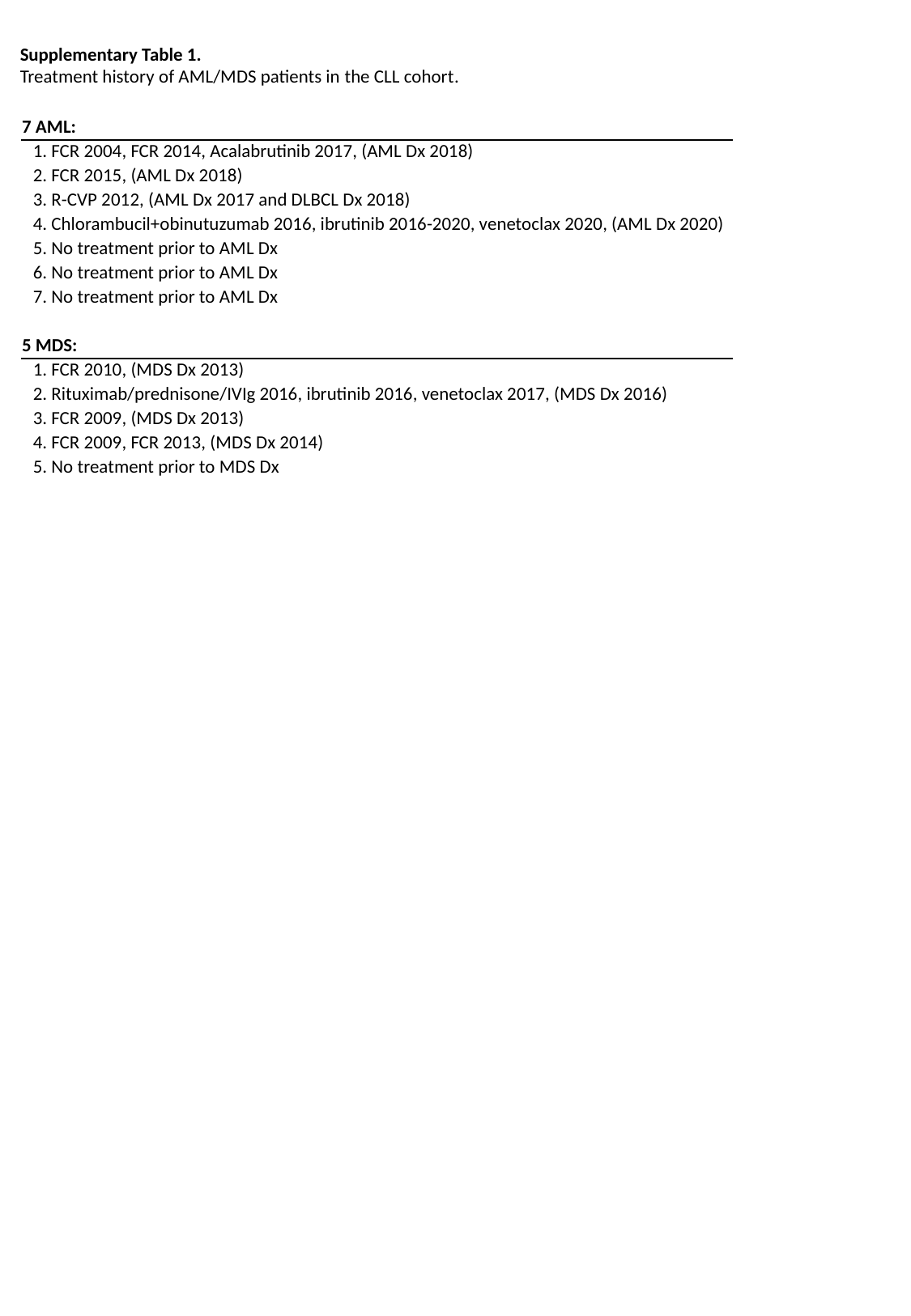

Supplementary Table 1.
Treatment history of AML/MDS patients in the CLL cohort.
| 7 AML: |
| --- |
| 1. FCR 2004, FCR 2014, Acalabrutinib 2017, (AML Dx 2018) |
| 2. FCR 2015, (AML Dx 2018) |
| 3. R-CVP 2012, (AML Dx 2017 and DLBCL Dx 2018) |
| 4. Chlorambucil+obinutuzumab 2016, ibrutinib 2016-2020, venetoclax 2020, (AML Dx 2020) |
| 5. No treatment prior to AML Dx |
| 6. No treatment prior to AML Dx |
| 7. No treatment prior to AML Dx |
| 5 MDS: |
| 1. FCR 2010, (MDS Dx 2013) |
| 2. Rituximab/prednisone/IVIg 2016, ibrutinib 2016, venetoclax 2017, (MDS Dx 2016) |
| 3. FCR 2009, (MDS Dx 2013) |
| 4. FCR 2009, FCR 2013, (MDS Dx 2014) |
| 5. No treatment prior to MDS Dx |

### Slide 2
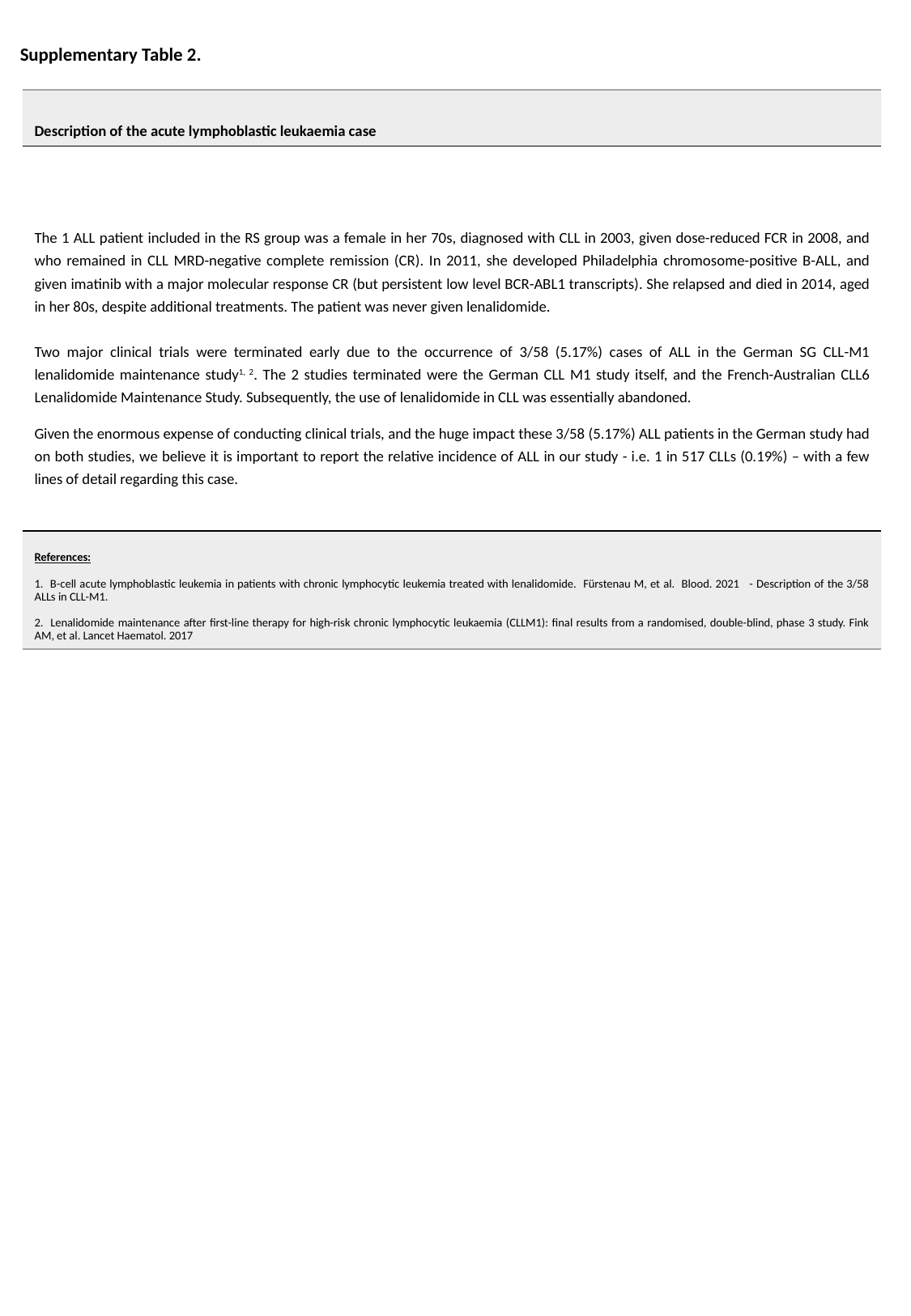

Supplementary Table 2.
| Description of the acute lymphoblastic leukaemia case |
| --- |
| The 1 ALL patient included in the RS group was a female in her 70s, diagnosed with CLL in 2003, given dose-reduced FCR in 2008, and who remained in CLL MRD-negative complete remission (CR). In 2011, she developed Philadelphia chromosome-positive B-ALL, and given imatinib with a major molecular response CR (but persistent low level BCR-ABL1 transcripts). She relapsed and died in 2014, aged in her 80s, despite additional treatments. The patient was never given lenalidomide. Two major clinical trials were terminated early due to the occurrence of 3/58 (5.17%) cases of ALL in the German SG CLL-M1 lenalidomide maintenance study^1, 2^. The 2 studies terminated were the German CLL M1 study itself, and the French-Australian CLL6 Lenalidomide Maintenance Study. Subsequently, the use of lenalidomide in CLL was essentially abandoned. Given the enormous expense of conducting clinical trials, and the huge impact these 3/58 (5.17%) ALL patients in the German study had on both studies, we believe it is important to report the relative incidence of ALL in our study - i.e. 1 in 517 CLLs (0.19%) – with a few lines of detail regarding this case. |
| References: 1. B-cell acute lymphoblastic leukemia in patients with chronic lymphocytic leukemia treated with lenalidomide. Fürstenau M, et al. Blood. 2021 - Description of the 3/58 ALLs in CLL-M1. 2. Lenalidomide maintenance after first-line therapy for high-risk chronic lymphocytic leukaemia (CLLM1): final results from a randomised, double-blind, phase 3 study. Fink AM, et al. Lancet Haematol. 2017 |

### Slide 3
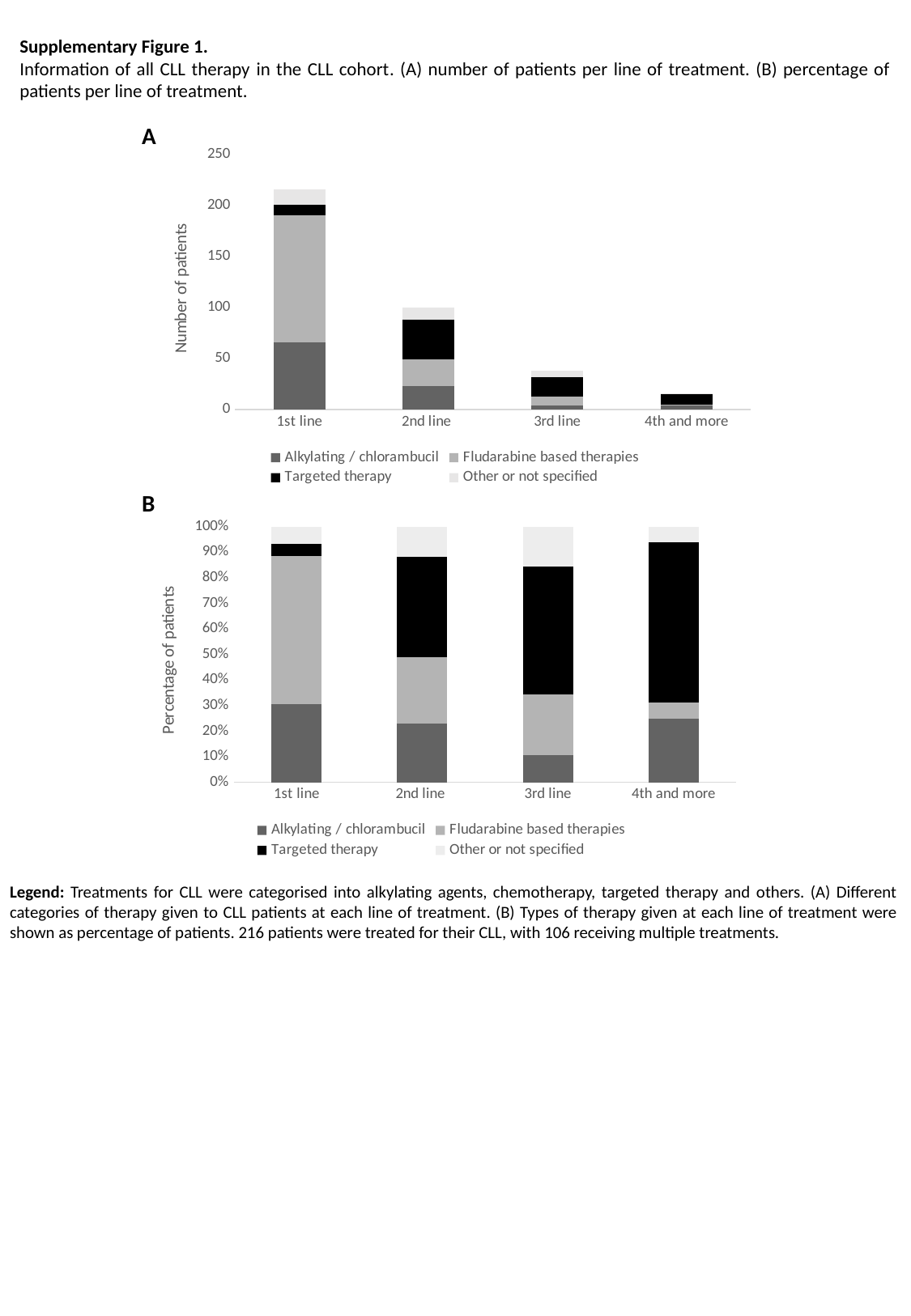

Supplementary Figure 1.
Information of all CLL therapy in the CLL cohort. (A) number of patients per line of treatment. (B) percentage of patients per line of treatment.
A
#### Chart
| Category | Alkylating / chlorambucil | Fludarabine based therapies | Targeted therapy | Other or not specified |
|---|---|---|---|---|
| 1st line | 66.0 | 125.0 | 10.0 | 15.0 |
| 2nd line | 23.0 | 26.0 | 39.0 | 12.0 |
| 3rd line | 4.0 | 9.0 | 19.0 | 6.0 |
| 4th and more | 4.0 | 1.0 | 10.0 | 1.0 |B
#### Chart
| Category | Alkylating / chlorambucil | Fludarabine based therapies | Targeted therapy | Other or not specified |
|---|---|---|---|---|
| 1st line | 0.3055555555555556 | 0.5787037037037037 | 0.046296296296296294 | 0.06944444444444445 |
| 2nd line | 0.23 | 0.26 | 0.39 | 0.12 |
| 3rd line | 0.10526315789473684 | 0.23684210526315788 | 0.5 | 0.15789473684210525 |
| 4th and more | 0.25 | 0.0625 | 0.625 | 0.0625 |Legend: Treatments for CLL were categorised into alkylating agents, chemotherapy, targeted therapy and others. (A) Different categories of therapy given to CLL patients at each line of treatment. (B) Types of therapy given at each line of treatment were shown as percentage of patients. 216 patients were treated for their CLL, with 106 receiving multiple treatments.

### Slide 4
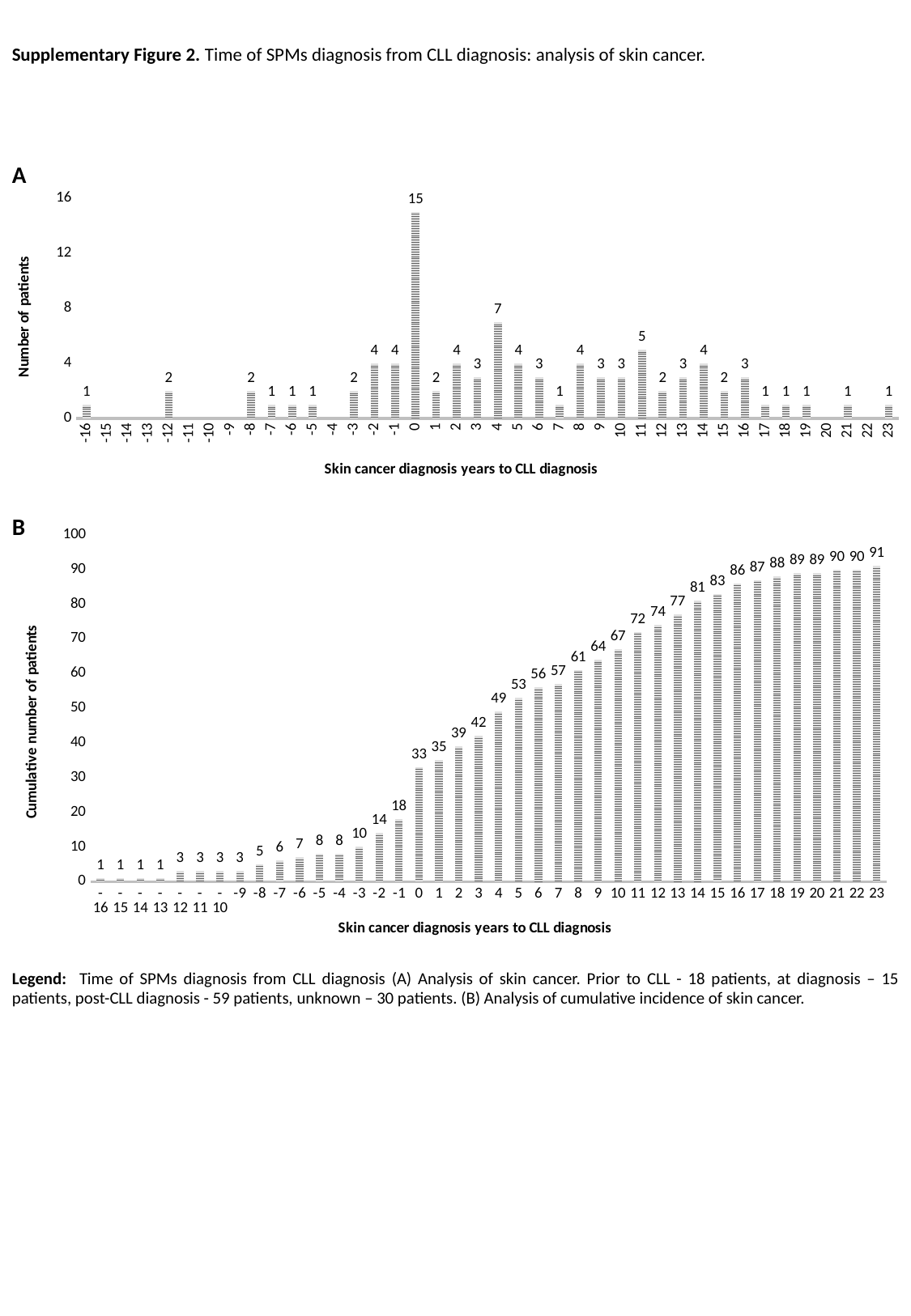

Supplementary Figure 2. Time of SPMs diagnosis from CLL diagnosis: analysis of skin cancer.
A
#### Chart
| Category | # of patients |
|---|---|
| -16 | 1.0 |
| -15 | None |
| -14 | None |
| -13 | None |
| -12 | 2.0 |
| -11 | None |
| -10 | None |
| -9 | None |
| -8 | 2.0 |
| -7 | 1.0 |
| -6 | 1.0 |
| -5 | 1.0 |
| -4 | None |
| -3 | 2.0 |
| -2 | 4.0 |
| -1 | 4.0 |
| 0 | 15.0 |
| 1 | 2.0 |
| 2 | 4.0 |
| 3 | 3.0 |
| 4 | 7.0 |
| 5 | 4.0 |
| 6 | 3.0 |
| 7 | 1.0 |
| 8 | 4.0 |
| 9 | 3.0 |
| 10 | 3.0 |
| 11 | 5.0 |
| 12 | 2.0 |
| 13 | 3.0 |
| 14 | 4.0 |
| 15 | 2.0 |
| 16 | 3.0 |
| 17 | 1.0 |
| 18 | 1.0 |
| 19 | 1.0 |
| 20 | None |
| 21 | 1.0 |
| 22 | None |
| 23 | 1.0 |
#### Chart
| Category | |
|---|---|
| -16 | 1.0 |
| -15 | 1.0 |
| -14 | 1.0 |
| -13 | 1.0 |
| -12 | 3.0 |
| -11 | 3.0 |
| -10 | 3.0 |
| -9 | 3.0 |
| -8 | 5.0 |
| -7 | 6.0 |
| -6 | 7.0 |
| -5 | 8.0 |
| -4 | 8.0 |
| -3 | 10.0 |
| -2 | 14.0 |
| -1 | 18.0 |
| 0 | 33.0 |
| 1 | 35.0 |
| 2 | 39.0 |
| 3 | 42.0 |
| 4 | 49.0 |
| 5 | 53.0 |
| 6 | 56.0 |
| 7 | 57.0 |
| 8 | 61.0 |
| 9 | 64.0 |
| 10 | 67.0 |
| 11 | 72.0 |
| 12 | 74.0 |
| 13 | 77.0 |
| 14 | 81.0 |
| 15 | 83.0 |
| 16 | 86.0 |
| 17 | 87.0 |
| 18 | 88.0 |
| 19 | 89.0 |
| 20 | 89.0 |
| 21 | 90.0 |
| 22 | 90.0 |
| 23 | 91.0 |B
Legend: Time of SPMs diagnosis from CLL diagnosis (A) Analysis of skin cancer. Prior to CLL - 18 patients, at diagnosis – 15 patients, post-CLL diagnosis - 59 patients, unknown – 30 patients. (B) Analysis of cumulative incidence of skin cancer.

### Slide 5
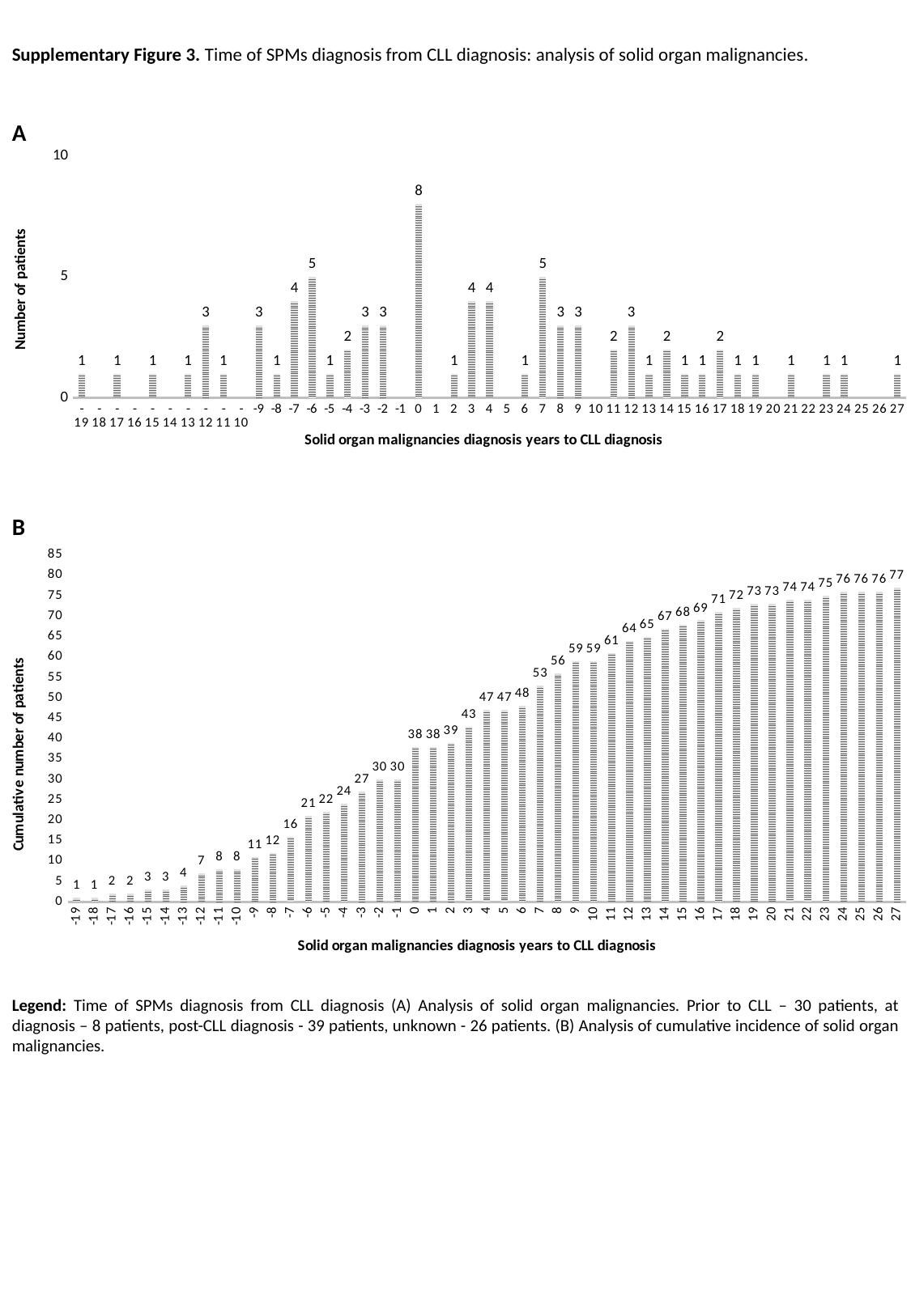

Supplementary Figure 3. Time of SPMs diagnosis from CLL diagnosis: analysis of solid organ malignancies.
A
#### Chart
| Category | # of patients |
|---|---|
| -19 | 1.0 |
| -18 | None |
| -17 | 1.0 |
| -16 | None |
| -15 | 1.0 |
| -14 | None |
| -13 | 1.0 |
| -12 | 3.0 |
| -11 | 1.0 |
| -10 | None |
| -9 | 3.0 |
| -8 | 1.0 |
| -7 | 4.0 |
| -6 | 5.0 |
| -5 | 1.0 |
| -4 | 2.0 |
| -3 | 3.0 |
| -2 | 3.0 |
| -1 | None |
| 0 | 8.0 |
| 1 | None |
| 2 | 1.0 |
| 3 | 4.0 |
| 4 | 4.0 |
| 5 | None |
| 6 | 1.0 |
| 7 | 5.0 |
| 8 | 3.0 |
| 9 | 3.0 |
| 10 | None |
| 11 | 2.0 |
| 12 | 3.0 |
| 13 | 1.0 |
| 14 | 2.0 |
| 15 | 1.0 |
| 16 | 1.0 |
| 17 | 2.0 |
| 18 | 1.0 |
| 19 | 1.0 |
| 20 | None |
| 21 | 1.0 |
| 22 | None |
| 23 | 1.0 |
| 24 | 1.0 |
| 25 | None |
| 26 | None |
| 27 | 1.0 |B
#### Chart
| Category | |
|---|---|
| -19 | 1.0 |
| -18 | 1.0 |
| -17 | 2.0 |
| -16 | 2.0 |
| -15 | 3.0 |
| -14 | 3.0 |
| -13 | 4.0 |
| -12 | 7.0 |
| -11 | 8.0 |
| -10 | 8.0 |
| -9 | 11.0 |
| -8 | 12.0 |
| -7 | 16.0 |
| -6 | 21.0 |
| -5 | 22.0 |
| -4 | 24.0 |
| -3 | 27.0 |
| -2 | 30.0 |
| -1 | 30.0 |
| 0 | 38.0 |
| 1 | 38.0 |
| 2 | 39.0 |
| 3 | 43.0 |
| 4 | 47.0 |
| 5 | 47.0 |
| 6 | 48.0 |
| 7 | 53.0 |
| 8 | 56.0 |
| 9 | 59.0 |
| 10 | 59.0 |
| 11 | 61.0 |
| 12 | 64.0 |
| 13 | 65.0 |
| 14 | 67.0 |
| 15 | 68.0 |
| 16 | 69.0 |
| 17 | 71.0 |
| 18 | 72.0 |
| 19 | 73.0 |
| 20 | 73.0 |
| 21 | 74.0 |
| 22 | 74.0 |
| 23 | 75.0 |
| 24 | 76.0 |
| 25 | 76.0 |
| 26 | 76.0 |
| 27 | 77.0 |Legend: Time of SPMs diagnosis from CLL diagnosis (A) Analysis of solid organ malignancies. Prior to CLL – 30 patients, at diagnosis – 8 patients, post-CLL diagnosis - 39 patients, unknown - 26 patients. (B) Analysis of cumulative incidence of solid organ malignancies.

### Slide 6
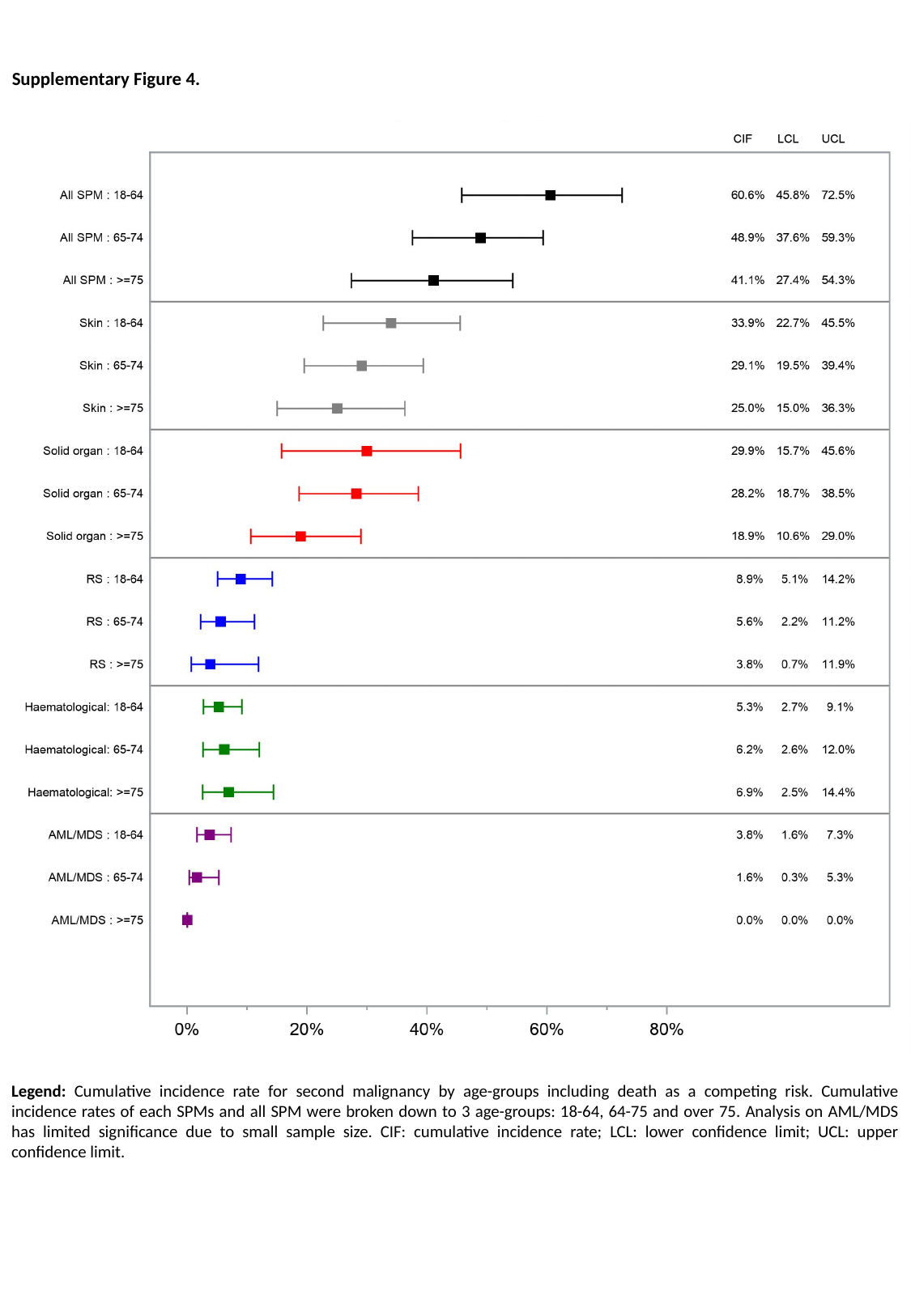

Supplementary Figure 4.
Legend: Cumulative incidence rate for second malignancy by age-groups including death as a competing risk. Cumulative incidence rates of each SPMs and all SPM were broken down to 3 age-groups: 18-64, 64-75 and over 75. Analysis on AML/MDS has limited significance due to small sample size. CIF: cumulative incidence rate; LCL: lower confidence limit; UCL: upper confidence limit.

### Slide 7
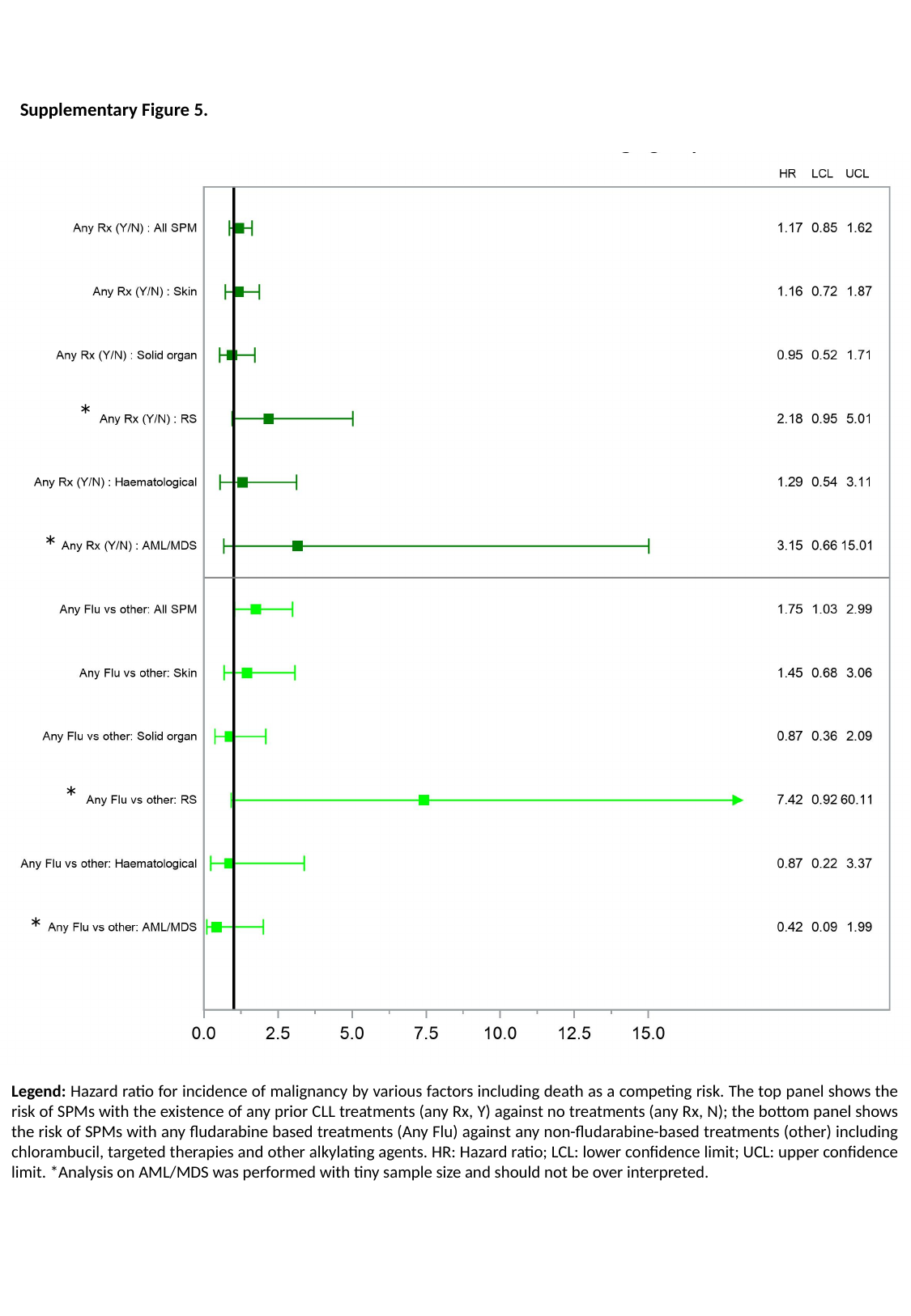

Supplementary Figure 5.
*
*
*
*
Legend: Hazard ratio for incidence of malignancy by various factors including death as a competing risk. The top panel shows the risk of SPMs with the existence of any prior CLL treatments (any Rx, Y) against no treatments (any Rx, N); the bottom panel shows the risk of SPMs with any fludarabine based treatments (Any Flu) against any non-fludarabine-based treatments (other) including chlorambucil, targeted therapies and other alkylating agents. HR: Hazard ratio; LCL: lower confidence limit; UCL: upper confidence limit. *Analysis on AML/MDS was performed with tiny sample size and should not be over interpreted.

### Slide 8
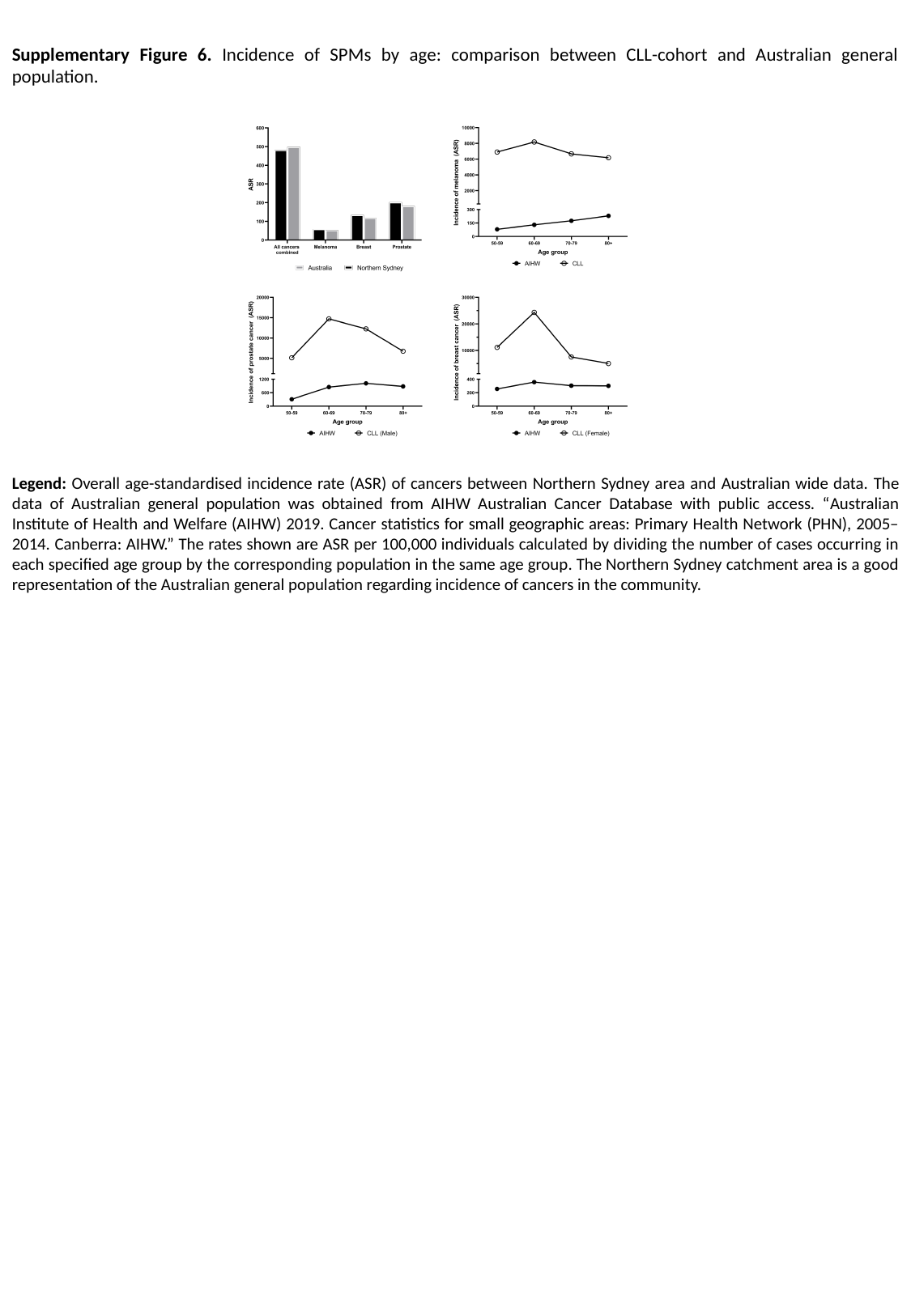

Supplementary Figure 6. Incidence of SPMs by age: comparison between CLL-cohort and Australian general population.
Legend: Overall age-standardised incidence rate (ASR) of cancers between Northern Sydney area and Australian wide data. The data of Australian general population was obtained from AIHW Australian Cancer Database with public access. “Australian Institute of Health and Welfare (AIHW) 2019. Cancer statistics for small geographic areas: Primary Health Network (PHN), 2005–2014. Canberra: AIHW.” The rates shown are ASR per 100,000 individuals calculated by dividing the number of cases occurring in each specified age group by the corresponding population in the same age group. The Northern Sydney catchment area is a good representation of the Australian general population regarding incidence of cancers in the community.
